# Seroprevalence and seroconversion rates to SARS-CoV-2 in interns, residents, and medical doctors in a University Hospital in Bogotá, Colombia

**DOI:** 10.1101/2020.09.15.20195313

**Authors:** Beatriz Ariza, Ximena Torres, Diana Salgado, Magda Cepeda, Carlos Gómez Restrepo, Julio Cesar Castellanos, Fernando Suárez-Obando, Adriana Cuellar, Claudia Cardozo, Juana Ángel, Manuel Antonio Franco

## Abstract

**Objectives:** To determine the prevalence of antibodies to SARS-CoV-2 and the incidence of seroconversion in the first month of follow-up among interns, residents, and medical doctors attending patients at a University Hospital, to explore for associations of seroprevalence and seroconversion with risk factors and symptoms compatible with COVID-19, and to explore the concordance of CLA, LFA, and ELFA.

**Design or methods:** We conducted a cross-sectional and a prospective study among medical doctors and medical trainees at Hospital Universitario San Ignacio in Bogota (Colombia) during June, July, and August to assess seroprevalence and seroconversion rates in this population was performed using CLA IgG for SARS-CoV-2. LFA IgG and IgM and ELFA IgM were also determined to explore concordance with CLA IgG.

**Results:** At baseline, 8 (2.28% 95%CI 1.16-4.43%) individuals were IgG positive for SARS-CoV-2 by CLA. At the end of the study, 21 (5.98% 95%CI 3.94-8.97%) individuals seroconverted by CLA IgG. In all, 29 individuals had IgG by CLA and of these 11 (3.13% 95%CI 1.76-5.52%) were asymptomatic. No associations with risk factors for infection were identified. CLA had moderate concordance with LFA IgG and ELFA, but minimal with LFA IgM.

**Conclusions:** Our report is one of the first in Latina America on seroprevalence and seroconversion rates in medical healthcare workers. It emphasizes the importance of avoiding focusing only on symptomatic individuals to screen this population for SARS-CoV-2 infection, since of all individuals that have evidence of previous infection many (37.93%) may be pre-symptomatic or asymptomatic and may contribute to infection/disease spread.

**Highlights:**

- Latin America was one of the most severely compromised regions of the world during the SARS-CoV-2 pandemic, between June and August 2020.
- Healthcare workers are at increased risk for COVID-19 and studies of seroprevalence and seroconversion rates in these subjects have not been published in the area.
- We conducted a cross-sectional and prospective study of medical doctors and medical trainees in a University Hosptial during June, July, and August 2020 to assess seroprevalence and seroconversion rates of SARS-CoV-2 in this population, using a Chemiluminescent assay (CLA).
- At baseline, 8 (2.28% 95%CI 1.16-4.43%) individuals were IgG positive for SARS-CoV-2 by CLA. At the end of the study, 21 (5.98% 95%CI 3.94-8.97%) individuals had seroconverted by CLA IgG.
- In all, 29 (8.26% 95%CI 5.81-11.61%) individuals had IgG for SARS-CoV-2 by CLA and of these 11 (3.13% 95%CI 1.76-5.52%) were asymptomatic.

## Introduction

During the SARS-CoV-2 pandemic healthcare workers (HCW) have been shown to have an increased risk of infection [1–6]. Studies in this population in many parts of the world have shown seroprevalences of between 2.4% and 45%, and in general above that of the general population and varing according to multiple factors [1–6]. In asymptomatic HCW, at the peak of the pandemic in England, a global seroprevalence rate of 24.4% was found [7]. Furthermore, individuals who retrospectively reported symptoms compatible with COVID-19 had a higher seroprevalence rate than those who did not report them, in this study and other studies [6,7]. Although retrospective reporting of symptoms may have evocation bias, these findings indicates that, in the context of COVID-19 a relationship can be established between retrospectively reported symptoms and seroprevalence. Seroconversion rates in HCW have been reported in fewer studies and varied between 20-44% in short term followup during high circulation of SARS-CoV-2 [1,8].

Latin-America is one of the most affected regions of the world by the pandemic [9], with peak cases occurring between July 20 and August 16 [10]. Although some studies from Latin-American countries evaluating serology in the general population [11,12] or schools have been published [13,14], to our knowledge only one study in an oncology unit in Brazil [15] has assessed seroprevalence in HCW. Our study was performed during a very active increase of SARS-CoV-2 infections in our country and city: during the five weeks of the study 248,205 new cases were identified in Colombia (109,505 *vs*. 357,710) and 93,907 of these were in Bogotá (34,131 *vs*. 128,038) (http://saludata.saludcapital.gov.co/osb/index.php/datos-de-salud/enfermedades-trasmisibles/covid19/ page consulted 08/19/20). Records from our hospital (Hospital Universitario San Ignacio (HUSI)) show that during June-August the adult intensive care unit (28 beds in June and 32 beds in July and August) was at full (100%) occupancy with presumed or confirmed COVID-19 patients.

The main purpose of this study was to determine the prevalence of antibodies, and the seroconversion rates to SARS-CoV-2 in a month of follow-up of interns, residents, and medical doctors of the School of Medicine of the Pontificia Universidad Javeriana attending patients at HUSI.

## Methods

### A) Study design

First, a cross-sectional study was conducted to determine the seroprevalence of SARS-CoV-2 infection in study population (interns, residents, and medical doctors that were treating patients at HUSI at the time of the study). Potential candidates were invited to participate by email. Potential participants who were not attending patients at HUSI in June and July and who were taking immunosuppressive drugs (chloroquine, corticosteroids, etc.) were excluded from the study (Figure 1). The remaining participants were asked to fill out a survey about risk factors and symptoms associated with COVID-19, history of previous diagnosis of COVID-19 confirmed by PCR or of clinically diagnosed COVID-19 supported by the presence of SARS-CoV-2 antibodies. The survey was designed in RedCap (Research Electronico Data capture [16]).

**Figure 1.**
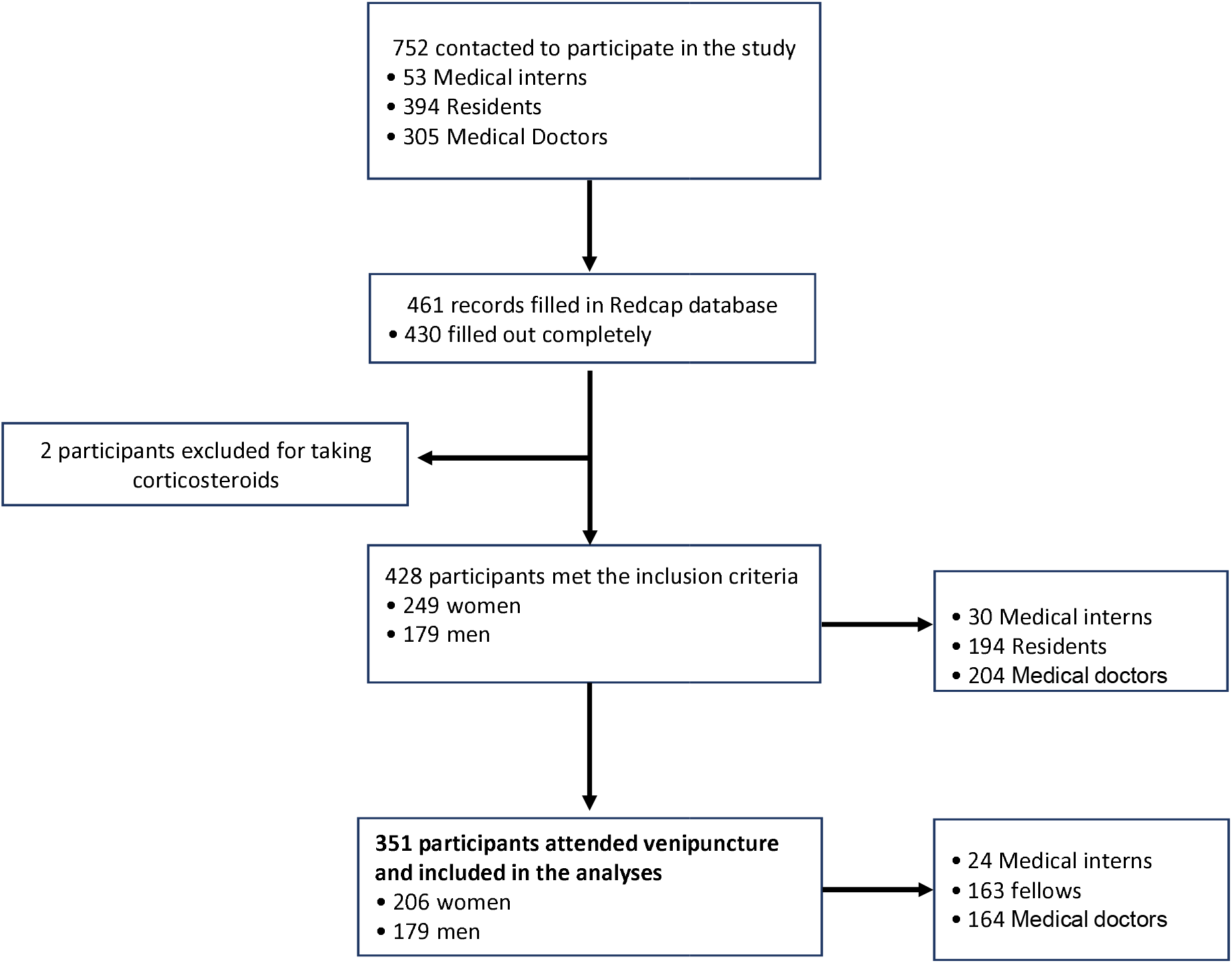
Study Flowchart

In a second step, a prospective study was conducted to determine the incidence of seroconversion two weeks and a month of follow-up after baseline visit among the seronegative individuals from the cross-sectional study.

As secondary objectives, we aimed to assess the relation between seropositivity either at baseline or during follow-up and risk factors and symptoms compatible with COVID-19. Finally, as an exploratory objective, we examined the concordance of CLA IgG as a tempative gold standard with the LFA IgG and IgM and ELFA IgM and concordance of the ELFA IgM and LFA IgM.

### B) Sampling and laboratory methods

At the HUSI’s clinical laboratory, individuals updated the survey of clinical symptoms compatible with COVID-19, signed an electronic informed consent, and donated 7 ml of venous blood.

#### Chemiluminescent assay (CLA)

SARS-CoV-2 IgG tests (Abbott Colombia) were performed on an Abbott Architect i1000 analyzer, following the manufacturer’s protocol. A single lot of positive and negative controls were run at the start of each batch of antibody testing. Samples with a signal-to-cutoff (S/CO) ratio greater than or equal to 1.4 were considered positive.

#### Lateral flow assays (LFA)

SARS-CoV-2 STANDARD Q COVID-19 IgM/IgG Duo Test kits (SD Biosensor, Gyeonggi-do, Korea) were performed following the manufacturer’s protocols. Positive results were determined by the appearance of a visible band in the designated area, simultaneously with an appropriate positive control band.

#### Enzyme linked fluorescence assay (ELFA)

The VIDAS Anti-SARS CoV-2 IgM two-step sandwich ELFA was performed on a VIDAS analyzer (BioMérieux, Marcy-l’Etoile, France). An index is calculated as the ratio between the relative fluorescence value measured in the sample and the relative fluorescence obtained for a calibrator (humanized recombinant anti-SARS CoV-2 IgM) and interpreted as negative (index<1) or positive (index≥1) [17].

All assays were validated with serum samples from PCR+/− individuals in our laboratory

### Ethical considerations

Our project complied with the legal and ethical guidelines contemplated in the Declaration of Helsinki of the World Medical Association, Fortaleza, Brazil, 2013. Likewise, it adheres to the ethical considerations outlined in articles 15 and 16 of Resolution No. 008430 of 1993 of the Ministry of Health and in Law 84 of 1989. The study and the informed consent form were approved by the ethics committee of School of Medicine of the Pontificia Universidad Javeriana and HUSI.

### Statistical analysis

The data was exported and analyzed in Stata 14. We conducted a descriptive analysis of the demographic characteristics of the study participants, according to the seropositivity. Continuous variables were described using median and interquartile range (percentiles 25th and 75th) and categorical variables were described using absolute and relative frequencies.

Second, we examined the relation between seropositivity either at baseline or during the follow-up and risk factors and symptoms compatible with COVID-19, we estimated the odds ratio and 95% confidence interval using logistic regression

Third, we assessed the concordance of CLA IgG, as a tempative gold standard, with LFA IgG and IgM and ELFA IgM, and the concordance of ELFA IgM and LFA IgM, using the Cohen’s kappa and the corresponding 95% confidence interval.

## Results

### Study population

Seven hundred and fifty-two (752) medical trainees or medical doctors from HUSI were invited to participate by email (Figure 1). Of these, 428 answered the baseline survey, and it was possible to arrange an appointment to bleed 351 of them (Figure 1). Six individuals reported a previous diagnosis of infection with SARS-CoV-2 confirmed by PCR (all but one with symptoms compatible with COVID-19) and two had been hospitalized with symptoms consistent with COVID-19 and positive SARS-CoV-2 antibodies, but their PCR had not identified SARS-CoV-2 (Tables 1 and 2 and Data not shown).

**Table 1.** Antibodies and symptoms of participants with at least one positive antibody results at baseline.

| Volunteer | Sample 1 |  |  |  |  | Sample 2 |  |  | Sample 3 |  |  |  |
| --- | --- | --- | --- | --- | --- | --- | --- | --- | --- | --- | --- | --- |
|  | Date | Symptoms* | IgM LFA | IgG LFA | IgG CLA | Date | Symptoms* | IgG CLA | Date | Symptoms* | IgG CLA |  |
| 1 | Jul 04 | NO | + | + | + | Jul 21 | NO | + | Aug 05 | NO | - | CLA positive at Baseline |
| 2 | Jul 03 | NO | + | + | + | Jul 17 | NO | + | Aug 01 | NO | + |  |
| 3 | Jun 25 | NO | + | + | + | Jul 09 | NO | + | Jul 23 | NO | + |  |
| 4 | Jun 30 | NO | - | - | + | Jul 16 | NO | + | Jul 30 | NO | + |  |
| 5 | Jul 01 | YES | + | + | + | Jul 15 | NO | + | Jul 29 | NO | + |  |
| 6 | Jul 04 | YES | + | + | + | Jul 18 | NO | + | Aug 01 | NO | + |  |
| 7 | Jul 02 | YES | + | + | + | Jul 16 | NO | + | Jul 30 | NO | + |  |
| 8 | Jun 30 | YES | - | - | + | Jul 14 | NO | + | Jul 28 | NO | + |  |
| 9 | Jun 27 | NO | - | + | - | Jul 11 | NO | - | Jul 25 | NO | - | LFA positive at Baseline |
| 10 | Jun 25 | NO | + | - | - | Jul 10 | NO | - | Jul 24 | NO | - |  |
| 11 | Jul 03 | NO | + | - | - | Jul 17 | NO | - | Jul 31 | NO | - |  |
| 12 | Jun 25 | NO | + | - | - | Jul 09 | NO | - | Jul 23 | NO | - |  |
| 13 | Jul 04 | NO | + | - | - | Jul 21 | NO | - | Aug 03 | NO | - |  |
| 14 | Jul 03 | NO | + | - | - | ND | ND | ND | Jul 31 | NO | - |  |
| 15 | Jun 25 | NO | + | - | - | Jul 10 | NO | - | Jul 24 | NO | - |  |
| 16 | Jun 30 | NO | + | - | - | Jul 14 | NO | - | Jul 28 | NO | - |  |
| 17 | Jun 26 | NO | + | - | - | Jul 10 | NO | - | Jul 24 | NO | - |  |
| 18 | Jun 30 | NO | + | - | - | Jul 14 | NO | - | Jul 28 | NO | - |  |
| 19 | Jul 04 | NO | + | - | - | Jul 21 | NO | - | Aug 04 | NO | - |  |
| 20 | Jun 26 | NO | + | - | - | Jul 10 | NO | - | Jul 24 | NO | - |  |
| 21 | Jun 26 | NO | + | - | - | Jul 10 | NO | - | Jul 24 | NO | - |  |
| 22 | Jul 03 | NO | + | - | - | Jul 17 | NO | - | Jul 31 | NO | - |  |
| 23 | Jun 30 | YES | + | - | - | Jul 15 | YES | - | ND | ND | ND |  |
| 24 | Jun 25 | NO | + | - | - | Jul 09 | NO | - | Jul 28 | NO | - |  |
| 25 | Jul 02 | NO | + | - | - | Jul 16 | NO | - | Aug 01 | NO | - |  |
| 26 | Jun 25 | YES | + | - | - | Jul 10 | NO | - | Jul 28 | NO | - |  |
| 27 | Jun 25 | NO | + | - | - | Jul 09 | NO | - | Jul 24 | NO | - |  |
| 28 | Jun 30 | NO | - | - | - | Jul 14 | YES | + | Jul 28 | NO | + | Seroconversion in CLA |
| 29 | Jun 26 | NO | - | - | - | Jul 10 | YES | + | Jul 25 | NO | + |  |
| 30 | Jun 26 | NO | - | - | - | Jul 13 | NO | + | Jul 24 | NO | + |  |
| 31 | Jun 30 | NO | - | - | - | Jul 15 | YES | + | Jul 30 | NO | + |  |
| 32 | Jun 25 | NO | - | - | - | Jul 13 | YES | + | Jul 23 | NO | + |  |
| 33 | Jul 03 | NO | - | - | - | Jul 17 | YES | + | Jul 31 | YES | + |  |
| 34 | Jul 03 | NO | - | - | - | Jul 17 | YES | + | Jul 31 | NO | + |  |
| 35 | Jun 26 | NO | - | - | - | Jul 15 | NO | + | Jul 24 | NO | + |  |
| 36 | Jul 03 | NO | - | - | - | Jul 17 | NO | + | Aug 03 | NO | + |  |
| 37 | Jun 25 | NO | - | - | - | Jul 09 | NO | - | Jul 27 | YES | + |  |
| 38 | Jul 03 | NO | - | - | - | Jul 17 | NO | - | Aug 10 | YES | + |  |
| 39 | Jun 30 | NO | - | - | - | Jul 14 | NO | - | Jul 30 | YES | + |  |
| 40 | Jul 03 | NO | - | - | - | Jul 17 | NO | - | Jul 31 | NO | + |  |
| 41 | Jun 25 | NO | - | - | - | Jul 09 | NO | - | Jul 28 | YES | + |  |
| 42 | Jul 03 | NO | - | - | - | Jul 17 | NO | - | Jul 31 | NO | + |  |
| 43 | Jul 01 | NO | - | - | - | Jul 15 | YES | - | Jul 31 | NO | + |  |
| 44 | Jun 30 | NO | - | - | - | Jul 17 | NO | - | Aug 08 | NO | + |  |
| 45 | Jun 26 | NO | - | - | - | ND | ND | ND | Jul 24 | NO | + |  |
| 46 | Jul 01 | NO | - | - | - | ND | ND | ND | Jul 29 | YES | + |  |
| 47 | Jul 03 | NO | - | - | - | ND | ND | ND | Aug 05 | YES | + |  |
| 48 | Jul 02 | YES | - | - | - | ND | ND | ND | Jul 31 | NO | + |  |

\* Symptoms before sample. Symptoms included cough, runny nose, fever, diarrhea, shortness of breath, sneeze, headache, odynophagia, dysgeusia, anosmia. ND; not done. CLA; Chemiluminescence assay. LFA; Lateral Flow Assay.

**Table 2.**
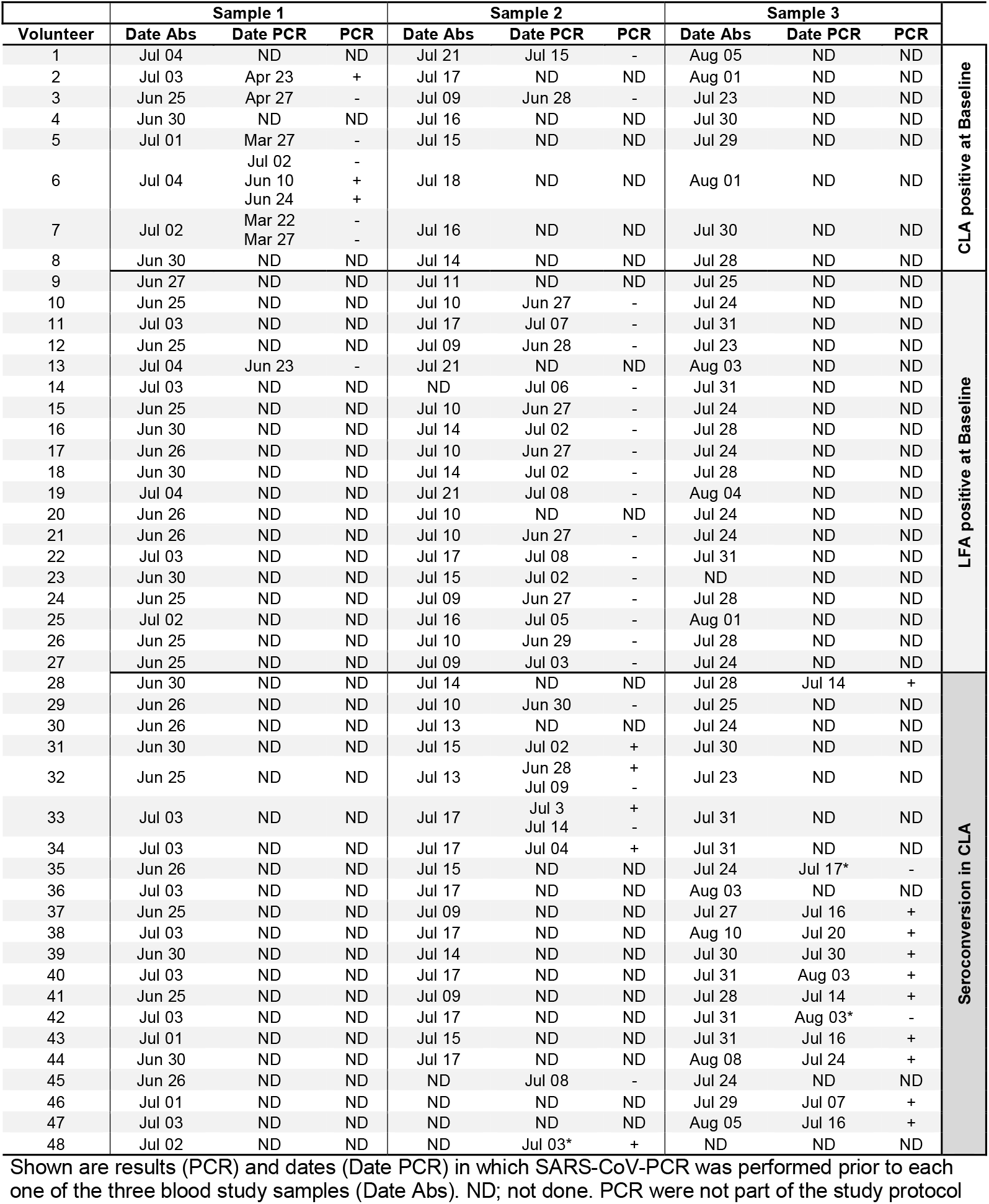
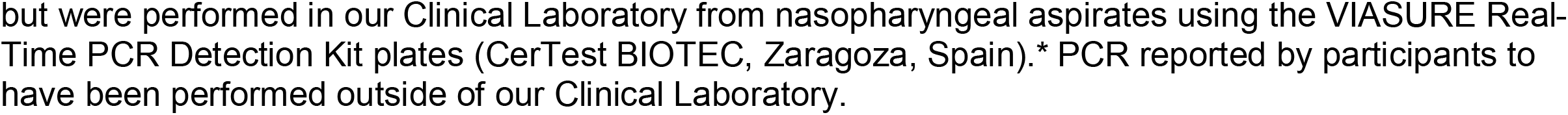
Date of blood sample and date and result of SARS-CoV-2 specific PCR performed in the study volunteers with at least one antibody positive results at baseline.

### Prevalence of SARS-CoV-2 antibodies in our cohort at baseline

Individuals in our cohort were bled at baseline between June 25 and the 4 of July. At baseline 8 (2.28% 95%CI 1.16-4.43%) individuals were SARS-CoV-2 IgG positive by CLA (Table 1 and Figure 2). For comparison, we also measured IgG and IgM antibodies by LFA (Table 1) and found that six individuals of the eight indivuals were also positive for IgM and IgG by LFA. Of these six individuals, one had COVID-19 compatible symptoms and a previous diagnosis of COVID-19 by PCR, two had previously been hospitalized with clinical diagnosis of COVID-19 (with negative PCR but positive serology), one had a positive PCR but had remained asymptomatic, and two without history of previous SARS-CoV-2 infection were also asymptomatic (Tables 1 and 2). Finally, one asymptomatic and one symptomatic individual were positive for IgG by CLA, but negative for LFA antibodies (Table 2). In addition, 18 individuals were only positive for SARS-CoV-2 LFA IgM and one was only positive for LFA IgG (Table 1). One of the 18 individuals that was only positive for IgM had a history of previous COVID-19 symptoms and a positive PCR before joining the study (Tables 1 and 2). The majority of individuals (16/18) with only a positive LFA IgM result and tested for SARS-CoV-2 PCR were negative for PCR at a date close to the date when the antibody sample was obtained (Table 2). Somewhat unexpectedly, three of the eight individuals that declared having a positive SARS-CoV-2 PCR previous to the start of the study were negative for all of the antibodies measured (Data not shown).

**Figure 2.**
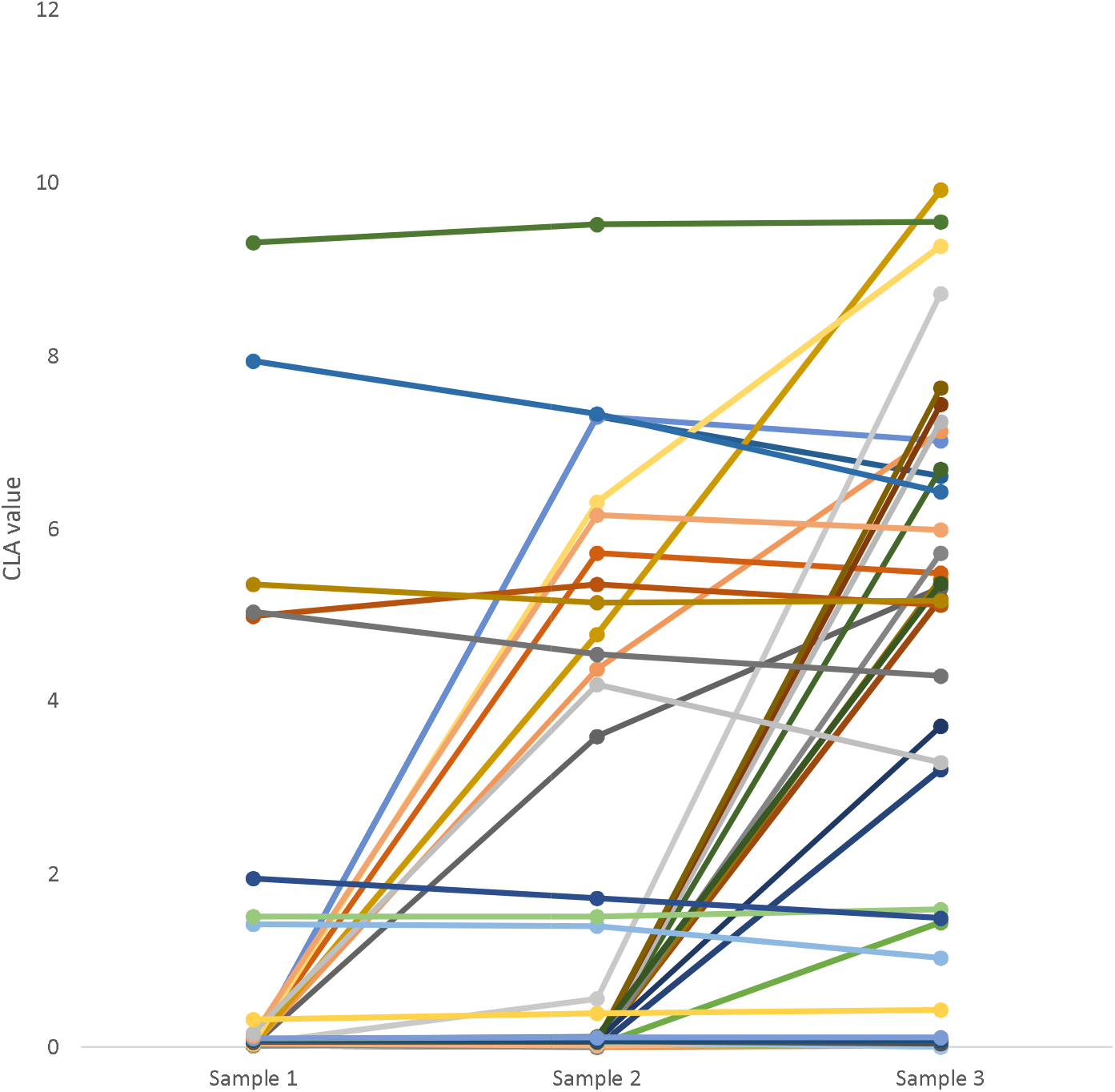
SARS-CoV-2 IgG CLA values at different times of blood sampling. Sample 1 was the baseline. Sample 2 was taken 15.1 days (95%CI 14.8-15.4)days after baseline. Sample 3 was taken 28.7 days (95%CI 28.3-29.0) after baseline. CLA; Chemiluminescence assay.

### Incidence of SARS-CoV-2 antibodies

A second and third blood samples was taken approximately two weeks (15.1 days 95%CI 14.8-15.4) and one month (28.7 days 95%CI 28.3-29.0) after baseline for each individual, from the 9th-21st of June and from June 23 to August 10, respectively.

Three hundred and thirty-five (335) of the original 351 (95.4%) individuals presented for the second bleeding. All eight initially positive individuals by CLA IgG remained positive (Table 1). Of the remaining 327 individuals, nine seroconverted in SARS-CoV-2 CLA IgG (2.75% 95%CI 1.45-5.14%, Table 1 and Figure 2). Three of these nine individuals were asymptomatic. None of the previously IgM positive individuals by LFA or the individual that only was IgG positive by LFA seroconverted by CLA IgG (Table 1) in the second bleed.

Three hundred and thirty-nine (339) of the original 351 (96.5%) individuals presented for the third bleeding. Seven of eight initially IgG positive individuals by CLA remained positive (Table 1), with one individual scoring marginally below the cutoff level of the assay (Figure 2). All nine individuals that seroconverted in IgG CLA in the second bleed remained positive and 12 new individuals (3.93% 95%CI 2.31-6.61%) seroconverted. Four of the twelve individuals that seroconverted in the last sample were asymptomatic (Table 1). None of the previously IgM positive individuals by LFA or the individual that only was IgG positive by LFA seroconverted by CLA (Table 1) in the third bleed. Altogether, we identified 21 individuals (5.98% 95%CI 3.94-8.97%) that seroconverted to SARS-CoV-2 IgG by CLA amongst our initial cohort of 351 individuals (Table 1 and Figure 2). Thus, adding the 21 individuals that seroconverted with the eight that had IgG by CLA at baseline, 29 individuals (8.26 95%CI 5.81-11.61%) had SARS-CoV-2 IgG by CLA and of these 11 (3.13% 95%CI 1.76-5.52%) were asymptomatic (Table 1).

### Associations of seroprevalence and seroconversion with risk factors and symptoms compatible with COVID-19

Demographic, infection risk factors, and prevalence of symptoms compatible with COVID-19 for this population are presented in Table 3 for individuals with or without a positive SARS-CoV-2 CLA test in the study. No risk factors were associated with seroprevalence or seroconversion to SARS-CoV-2.

**Table 3.** Comparison of demographics and risk factors of CLA IgG negative and positive patients

| Characteristic | Negative<br>(n = 322) | Positive<br>(n = 29) | OR (95% CI) |
| --- | --- | --- | --- |
| *Demographics* |  |  |  |
| Age, years; Median (IQR) | 31.5 (27.5 - 38.6) | 29.4 (26.9 - 37) | 0.98 (0.94 - 1.03) |
| Sex; n (%) |  |  |  |
| Women | 192 (59.6) | 14 (48.3) | Reference |
| Men | 130 (40.4) | 15 (51.7) | 1.58 (0.74 - 3.39) |
| Mode of transport; n (%) |  |  |  |
| Public transport | 7 (2.2) | 1 (3.4) | Reference |
| Car/moto | 256 (79.5) | 21 (72.4) | 0.57 (0.07 - 4.89) |
| Walking/Bycicle | 59 (18.3) | 7 (24.1) | 0.83 (0.09 - 7.78) |
| Service; n (%) |  |  |  |
| Emergencies | 66 (20.5) | 8 (27.6) | Reference |
| ICU | 11 (3.4) | 1 (3.4) | 0.75 (0.09 - 6.6) |
| Outpatient consultation | 60 (18.6) | 5 (17.2) | 0.69 (0.21 - 2.22) |
| Other | 185 (57.5) | 15 (51.7) | 0.67 (0.27 - 1.65) |
| Occupation; n (%) |  |  |  |
| Healthcare worker in training | 167 (51.9) | 20 (69) | Reference |
| Healthcare worker | 155 (48.1) | 9 (31) | 0.48 (0.21 - 1.1) |
| *Risk Factors* |  |  |  |
| Obesity; n (%) |  |  |  |
| No | 306 (95) | 27 (93.1) | Reference |
| Yes | 16 (5) | 2 (6.9) | 1.42 (0.31 - 6.49) |
| Smoking behavior; n (%) |  |  |  |
| No | 301 (93.5) | 26 (89.7) | Reference |
| Yes | 21 (6.5) | 3 (10.3) | 1.65 (0.46 - 5.91) |
| Diabetes diagnosis; n (%) |  |  |  |
| No | 319 (99.1) | 28 (96.6) | Reference |
| Yes | 3 (0.9) | 1 (3.4) | 3.8 (0.38 - 37.72) |
| Hypertension diagnosis; n (%) |  |  |  |
| No | 306 (95) | 26 (89.7) | Reference |
| Yes | 16 (5) | 3 (10.3) | 2.21 (0.6 - 8.07) |
| Symptoms before recruitment; n |  |  |  |
| (%) |  |  |  |
| No | 255 (79.2) | 24 (82.8) | Reference |
| Yes | 67 (20.8) | 5 (17.2) | 0.79 (0.29 - 2.16) |
| *COVID-19 exposure* |  |  |  |
| Close contact with COVID-19 patients; n (%) |  |  |  |
| No | 76 (23.6) | 7 (24.1) | Reference |
| Yes | 180 (55.9) | 17 (58.6) | 1.03 (0.41 - 2.57) |
| Not known | 66 (20.5) | 5 (17.2) | 0.82 (0.25 - 2.71) |
| Contact with body fluids; n (%) |  |  |  |
| No | 194 (60.2) | 13 (44.8) | Reference |
| Yes | 107 (33.2) | 13 (44.8) | 1.81 (0.81 - 4.05) |
| Not known | 21 (6.5) | 3 (10.3) | 2.13 (0.56 - 8.09) |
| Use of Personal Protection Elements; n (%) |  |  |  |
| No | 1 (0.3) | 0 (0) | Reference |
| Yes, complete per protocol | 294 (91.3) | 27 (93.1) | 1.24 (0.28 - 5.5) |
| Yes, incomplete | 27 (8.4) | 2 (6.9) | - |

### Concordance of the antibody assays

To further evaluate concordance of the LFA and CLA assays and to extend this analysis to ELFA IgM, thawed samples from the first bleed were tested by ELFA and thawed samples from bleeds 2 and 3 were tested by ELFA IgM and LFA IgG and IgM. Concordance of CLA IgG with LFA IgG and ELFA IgG was moderate and with LFA IgM, minimal (Table 4) [18]. The ELFA IgM and LFA IgM also had minimal concordance (Table 5).

**Table 4.**
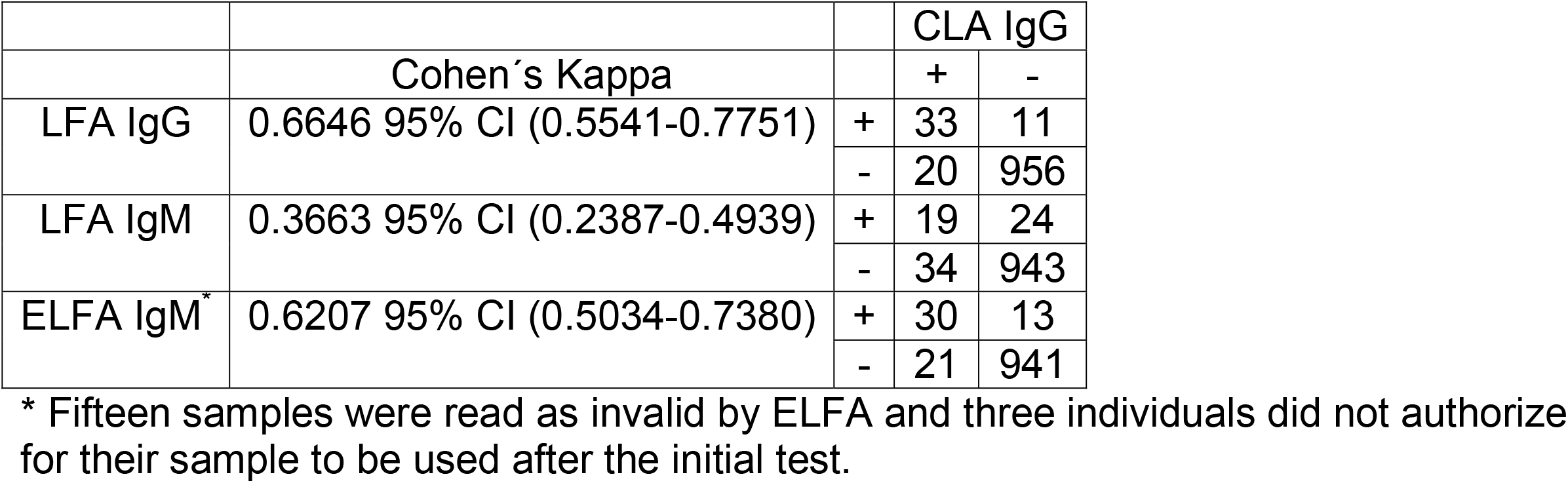
Concordance between CLA IgG and LFA IgG, LFA IgM, and ELFA IgM

**Table 5.**
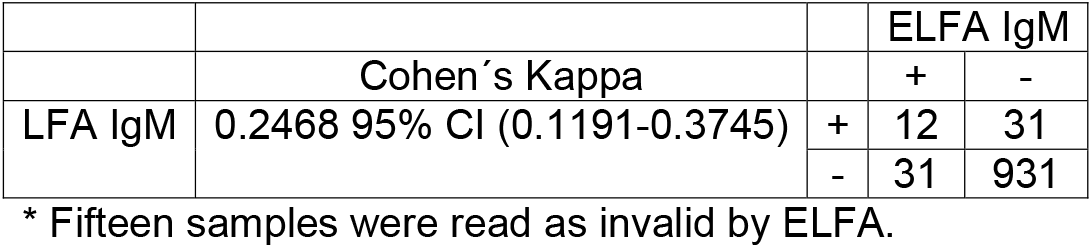
Concordance between ELFA IgM and LFA IgM

## Discussion

We have performed one of the first SARS-CoV-2 seroprevalence/seroconversion rate studies in Latin-America and found that at baseline 2.28% of HCW were IgG positive by CLA (Table 1). At the end of the study, 5.98% of individuals had seroconverted by CLA

IgG and, in all, 29 individuals (8.26%) had SARS-CoV-2 IgG by CLA, of which 11 (3.13%) were asymptomatic (Table 1). No associations between seroprevalence/seroconversion in CLA and risk factors for infection were identified. Concordance of CLA IgG with LFA IgG and ELFA IgG was moderate and with LFA IgM, minimal (Table 4). The ELFA IgM and LFA IgM also had minimal concordance (Table 5).

The levels of seroprevalence for CLA IgG (2.28%) at the beginning of the study and of seroconversion to this antibody (5.98%) are comparable to those reported in other studies and, overall, higher than those observed in the general population [1–6]. In a comparable study in England that followed 200 front line HCW for two weeks, they found that 20% of them seroconverted during the study, but 25% were already seropositive at the beginning of the study [1]. Most likely, the higher numbers in the English study compared with our study are due to the differences in the populations studied (front line workers vs a mixed population of medical doctors).

One of the main findings of our study, is the relatively high numbers (3.18%) of asymptomatic individuals positive for IgG by CLA (Table 1). This number is very close to the number of asymptomatic HCW detected by screening with PCR in nasofaringeal swabs (3%) [19] or saliva (2,6%) [20] in England. Although it is incompletely clear how much pre-symptomatic and asymptomatic individuals contribute to virus spread, focusing only on stopping symptomatic individuals is insufficient to control the spread of the virus [21,22]. None-invasive rapid screening strategies for SARS-CoV-2 infection are needed to evaluate symptomatic and asymptomatic HCW.

The lack of association between risk factors and SARS-CoV-2 seroprevalence/seroconversion in CLA (Table 3) may be explained because some of the risk factors evaluated (obesity, diabetes, hypertension, and smoking) can be risk factors for disease and not infection. Moreover, most of the participants used PPE and followed biosafety recommendations (Table 3).

Our results seem comparable to previous studies in which the CLA test that we used showed a sensitivity and specificity close to 100% when compared with PCR +/− samples [23–25], while the LFA [26] and the ELFA [17] appear to be less sensitive and specific. The LFA IgG seems to have missed 20 samples positive by CLA IgG (Tabel 4), and all but one of the 11 samples positive by LFA IgG but negative by CLA IgG were only positive for this antibody, suggesting they may be false positives. The minimal concordance of the LFA IgM with other assays can probably be explained because of a high level of false positives: at baseline most (18, 5.13% 95%CI 3.27-7.96) of the individuals that had any positive antibody were positive for LFA IgM only (Table 1). However, none of these individuals had a positive PCR at or close to the time when the sample was taken (Table 3). With few exceptions, they did not present with COVID-19 compatible symptoms (Table 1) and none of them seroconverted to IgG by CLA on follow-up. These results are consistent with the hypothesis that most, if not all, of these results are false-positive results. This hypothesis is in agreement with the validation performed by our National Institute of Health that reports that the IgM LFA assay may have 4% false positives defined using serums from prepandemic individuals (https://www.ins.gov.co/Pruebas_Rapidas/4.%20Informe%20de%20validaci%C3%B3n%20PR%20SD%20Biosensor.pdf page consulted August 25).

Our study may have a sampling bias. Independent data from our hospital indicate that up to the 15th of June 13 medical doctors and 7 residents from approximately 800 individuals had been diagnosed with COVID-19 (with PCR or clinical symptoms/serology). By the 15^th^ of August, these numbers had increased to 44 medical doctors and 55 residents diagnosed with COVID-19. These numbers are higher than what we found in our population an suggest that our values of seroprevalence and seroconversion may be underestimated. A probable explanation for this is that volunteers with COVID-19 were isolated at the time of sampling and were unable to participate in the study or that having been previously tested were uninterested in participating.

## Data Availability

All study data is available upon request

## Statements

## Acknowledgement

We want to thank the volunteers that participated and donated blood for this study. We want to thank Dr Pablo Aschner for careful reading and suggestion for the project and Madga Alba and Jenny Severiche for help with the process of recruiting volunteer.

## Ethical Statement

This study was approved by the Ethics Committee of the School of Medicine of Pontificia Universidad Javeriana Bogotá, Colombia and was done in accordance with

The Code of Ethics of the World Medical Association (Declaration of Helsinki). An informed consent form, approved by the Ethics Committee of HUSI and the Pontificia Universidad Javeriana School of Medicine, was signed by all volunteers.

## Author Contributions

Juana Angel AND Manuel Franco: Conceptualization; Data curation; Formal analysis; Funding acquisition; Project administration; Resources; Supervision; Writing - original draft; Writing - review & editing.

Ximena Torres and Diana Salgado: Data curation; Investigation; Writing - review & editing.

Beatriz Ayala: Conceptualization; Project administration; Investigation; Supervision; Writing - review & editing.

Claudia Cardozo: Conceptualization; Resources; Writing - review & editing.

Magda Cepeda: Conceptualization; Data curation; Software; Formal analysis; Writing - review & editing.

Adriana Cuellar, Fernando Suarez, Carlos Gómez-Restrepo, Julio Cesar Castellanos: Conceptualization; Writing - review & editing.

